# Association of Adenosine Deaminase (ADA) +22 G->A (rs73598374) Genotype in Latino Adult Obese

**DOI:** 10.1101/2021.11.11.21266231

**Authors:** Frank Martiniuk, Kam-Meng Tchou-Wong, Angela Chen, Alexander H. Wong, Esme Cribb, Gilbert Smolenski, Kyle Yocum, Anna Barangan, Kahmun Lo, Melissa Watt, Ian Fagan, Arthur Nadas, Chelsea O’Koren, Fritz Francois

## Abstract

Obesity is a health burden that currently affects over 13% of the global adult population, consisting of over 650 million adults. Obesity is evaluated based on a body mass index (BMI) scale, which is calculated as weight in kilograms divided by the square of height in meters. Adults with a BMI greater than 30kg/m^2^ are considered to be obese while adults with a BMI greater than 40kg/m^2^ are considered morbidly obese. Adenosine deaminase (ADA) is a polymorphic enzyme that plays an important role in both immune functions and the regulation of intracellular and extracellular concentrations of adenosine. Three alleles of the *ADA* gene (*ADA*^*1*^, *ADA*^*2*^, *ADA*^*6*^) are associated with type 1 diabetes mellitus (DM1). *ADA*^*2*/*6*^ may increase susceptibility to DM1 in females. Given the evidence linking obesity with DM1, we wanted to determine whether a correlation exists between *ADA*^*2*^ allele and obesity. The *ADA*^*2*^ (+22 G->A, rs73598374) SNP changes the amino acid at position 8 from aspartic acid (Asp8)(D) to asparagine (Asn)(N). In this study, we present significant evidence of association between the *ADA*^*2*^ allele and obesity or BMIs greater than 25 in the Latino adult but not in the Caucasian population and therefore, may be a risk for obesity and its complications such as DM1.

## INTRODUCTION

Obesity is a health burden that currently affects over 13% of the global adult population in 2016, consisting of over 650 million obese adults with more than 1.9 billion adults-18 years and older were overweight. Over 340 million children and adolescents aged 5-19 were overweight or obese [www.who.int/news-room/fact-sheets/detail/obesity-and-overweight][1,2]. It causes multiple heath problems and serious complications, such as coronary heart disease, cardiovascular arrests, and risk of diabetes [3, 4]. In the United States, the non-diagnosed diabetes mellitus type 2 (DMT2) has a prevalence as high as 6.9% [5]. By standard procedure, obesity is evaluated based on a body mass index (BMI) scale, which is calculated as weight in kilograms divided by the square of height in meters. BMI has been studied extensively due to the health effects of obesity and its relative ease of measurement. Excessive weight increases the risk of type II diabetes, coronary artery disease and hypertension and is associated with many forms of cancer along with adverse social and psychological consequences. The fundamental cause of obesity and overweight is an energy imbalance between calories consumed and calories expended. Globally, there has been an increased intake of energy-dense foods that are high in fat and sugars and an increase in physical inactivity due to the increasingly sedentary nature of many forms of work, changing modes of transportation and increasing urbanization.

Adults with a BMI greater than 30kg/m^2^ are considered to be obese while adults with a BMI greater than 40kg/m^2^ are considered morbidly obese [1]. Adults with a BMI between 25kg/m^2^ to 29.9kg/m^2^ are considered overweight and a normal BMI ranges from 18.5kg/m^2^ to 24.9kg/m^2^ [6]. Women are more likely to be obese, while men are more likely to be overweight [6]. Within the United States, 25% of children are overweight and 11% are considered obese [7]. Dehghan et al. [7] found that 70% of obese adolescents grew up to be obese adults. Currently, at least 18 genetic loci have been identified, each of which consist of several single nucleotide polymorphism (SNP) that are involved in the pathogenetic complexity of diabetes [4, 8]. Obesity can be caused by physical inactivity, a poor diet or both in combination. Block [9] found that people in the United States consumed about 25% of their total calories from nutrient-poor foods. Further, obesity is associated with diabetes mellitus, elevated levels of triglycerides and low density lipoprotein cholesterol (LDL-C) and lower levels of high density lipoprotein cholesterol (HDL-C) than people of average weight. Elevated levels of triglycerides and LDL-C and low levels of HDL-C are associated with increased risk for cardiovascular disease [3]. Adenosine deaminase (ADA) is a polymorphic enzyme that plays an important role in both immune functions and the regulation of intracellular and extracellular concentrations of adenosine. Deficiency of ADA results in severe combined immunodeficiency (SCID) presenting in infancy and usually resulting in early death [10-14]. There is an association between type 1 diabetes mellitus (DM1) and three alleles (*ADA*^*1*^, *ADA*^*2*^, *ADA*^*6*^) of the *ADA* gene. *ADA*^*2*/*6*^ alleles may increase susceptibility to DM1 in females [15]. The role of ADA has been implicated as an important regulator of insulin action and in the pathogenesis of diabetes mellitus. Increased serum levels of ADA and its correlations with glycemic control have been reported in diabetic patients [5,6]. Animal studies suggest that a genomic region of *ADA* modifies the rate at which sleep need accumulates during wakefulness [16]. The current study was conducted to determine whether a correlation exists between *ADA*^*2*^ allele and obesity. Finding a correlation between the *ADA2* allele and obesity will assist in explaining the underlying genetic contributor to obesity. Uncovering such a correlation would provide doctors with a means of testing for genetic predisposition to obesity and taking aggressive measures to overcome hereditary tendencies. The *ADA*^*2*^ (+22 G->A, exon 1) SNP changes the amino acid at position 8 from aspartic acid (GAC, Asp8)(D) to asparagine (AAC, Asn)(N). The negatively charged aspartic acid to the polar uncharged asparagine can affect the tertiary and secondary structures as well as the folding of the enzyme that affects the affinity of the ligand to the active site. *ADA*^*2*^ has 20% to 30% less enzyme activity than the *ADA*^*1*^ allozyme. Bottini et al. [17] studied 273 subjects with non-insulin-dependent diabetes mellitus (NIDDM) from Penne, Italy. A low proportion of the *ADA*^*2*^ allele was observed in NIDDM subjects with a BMI of 25kg/m^2^ or less and a high proportion of this allele is observed in NIDDM patients with a BMI higher than 34kg/m^2^. In the intermediate BMI class, the proportion of *ADA*^*2*^ allele does not differ significantly from that of normal subjects. No significant effect on the relation between ADA and BMI has been observed for sex, age, age at onset, therapy with insulin and dyslipidemia. Jadhav and Jain [18] showed the serum ADA activity was significantly increased in overweight and obese subjects and as well as in combined overweight and obese group as compared to control in overweight and obese Indian subjects. *ADA*^*2*^ is associated with duration, depth of deep sleep and important source of variation in sleep homeostasis in humans, through modulation of specific components of the sleep EEG [19-21]. Kurtul et al. [22] found significant increased serum ADA activity in 50 obese individuals from Turkey.

In this study, we wanted to determine whether a correlation exists between *ADA*^*2*^ allele and obesity. The *ADA*^*2*^ (+22 G->A, rs73598374) SNP changes the amino acid at position 8 from aspartic acid (Asp8)(D) to asparagine (Asn)(N). We present significant evidence of association between *ADA*^*2*^ allele and obesity or BMIs greater than 25kg/m^2^ in the Latino adult, but not in the Caucasian population and therefore, may be a risk for obesity.

## MATERIALS and METHODS

DNA was isolated from peripheral blood white blood cells using standard proteinase K-SDS digestion; phenol/chloroform extraction and ethanol precipitation methods [23] from randomly selected 229 individuals where age, sex, height and ethnic/racial group were recorded. PCR was utilized to amplify the *ADA* gene. The PCR reaction included 20 μ L of water, 10.0 μ L of the template DNA, 10.0 μ L of J buffer, 5.0 μ L of dimethylsulfoxide (DMSO), 4.0 μ L of 2.5 mM deoxyribonucleotide triphosphates (dNTPs), 0.5 μ L of Taq polymerase, 1 μ L of each outer primer (0.2 ug/ L). The cycling times were: disassociation at 94°C for one minute; annealing at 60°C for one minute; extension at 72°C for one minute for 45 cycles followed by 72 °C for 10 minutes. The primers are: 5’-CCGGCCCGTTAAGAAGAGCGTGGCCGGCCG-3’ and 5’-CTAAGC CGAAGGAAGAACTCGCCTGCAGGA-3’. Ten uL of PCR amplicons were electrophoresed on 2.5% agarose gel containing ethidium bromide. The gels were photographed with a FB-PDC-34 photo-documentation camera (Thermo-Fisher Scientific-Springfield, NJ). Five to 15 μ L of PCR amplicon was digested in a final volume of 40 uL containing 4 μ L of 10x C buffer, 1.0 μ L of BSA and 1 μ L of Taq1 at 657#x00B0;C for two hours. Samples were electrophoresed on 2.5% agarose gel at ∼60 volts for two to three hours and photographed.

The Prism4 statistical software package was used for statistical analysis. The SNP genotype percentage was compared between groups using the Pearson Chi-square test or Fisher**’**s exact method. P values less than 0.05 were considered statistically significant.

## RESULTS

Our 229 adult cohort consisted of 46% male and 54% female with 73 patients having a BMI of <25 and 156 patients having a BMI of >25. Thirty-three were White, 150 were Latino and 46 were Asian or of unknown ethnic origins (Table 1). We genotyped DNA from the 229 patients for the *ADA*^*2*^ allele (+22) of a G (*ADA*^*1*^) to A (*ADA*^*2*^) coding for Asp^8^ by restriction endonuclease TaqI digestion of PCR amplicons (Figure 1). TaqI digestion detects the G allele (*ADA*^*1*^)(TC**<u>G/A</u>**AC; TCGA = TaqI). The patients were sorted into four BMI groups: <25, 25-30, 30.1-35 and 35+ plus White and Latino ethnic subgroups (Table 1). Combining all patients with BMIs <25 (n = 63) vs. >25 (n = 156), we found a significant difference between the two groups for the frequency of the *ADA*^*2*^ allele (p = ≤ 0.0157). However, if we separate the White and Latino ethnic groups, we found that the significant difference in the frequency of the *ADA*^*2*^ allele in the Latino population and not the White population. For the White population, there was no significant difference between the BMIs <25 and the BMIs >25 for the frequency of the *ADA*^*2*^ allele (p = ≥ 0.05). For the Latino group, there was a significant difference between the two BMI groups for the frequency of the *ADA*^*2*^ allele (p = ≤ 0.0251). No additional differences were found if sorted by male and female.

**Table 1.**
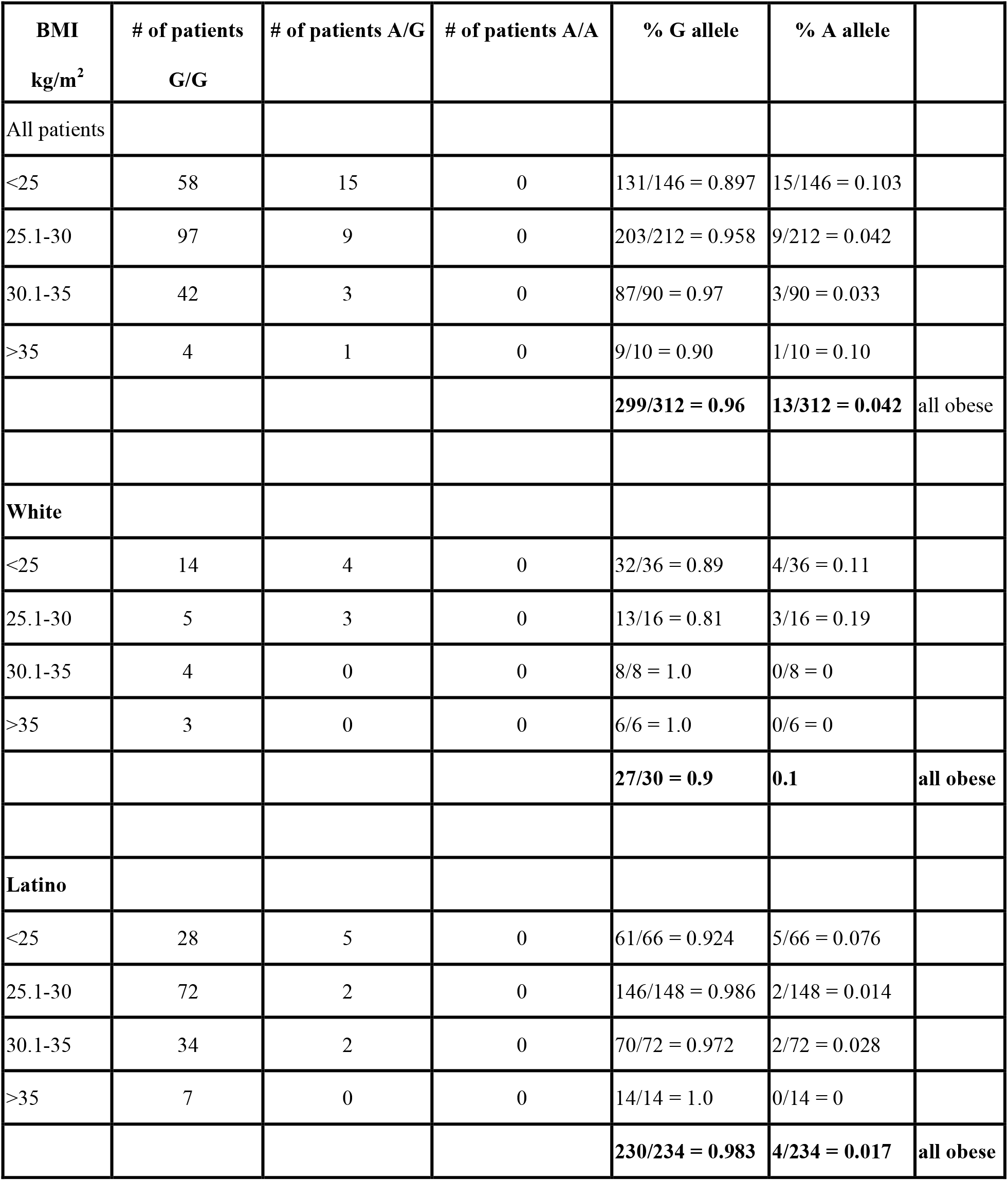
*ADA* genotypes in obese patients.

**Figure 1.**
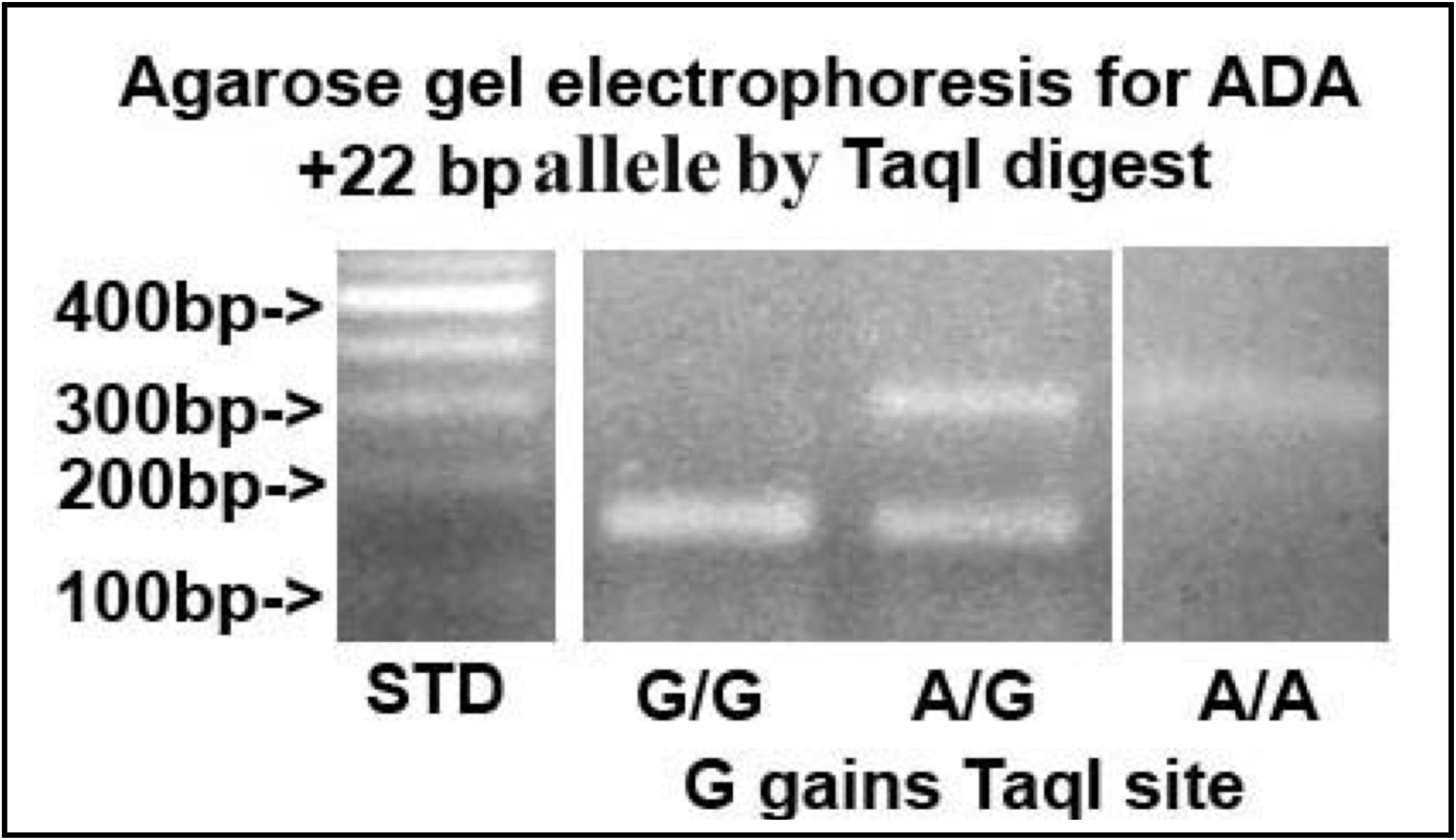
Composite photograph of agarose gel electrophoresis of human *ADA* PCR amplicons digested with TaqI. TaqI detects the allele containing a G (*ADA*^*1*^) that creates a TaqI restriction endonuclease site. Lane 1 on left shows 100 bp DNA standard ladder. Lane 2 from left shows DNA from an homozygous G/G sample, lane 3 from left shows DNA from an heterozygous G/A and lane 4 from the left shows DNA from a homnozygous A/A.

## DISCUSSION

Obesity prevalence is currently increasing due to various factors, including hypercaloric alimentation, decreased physical activity, social status and genetic variants [24]. The genotyping of 16 SNPs from published studies has shown that variants of 11 genes (*TCF7L2, PPARG, FTO, KCNJ11, Notch2, WFS1, CDKAL1, IGF2BP2, SLC30A8, JAZF1* and *HHEX*) are significantly associated with the risk of DMT2 and therefore, the risk of obesity. As a result, the association of genetic variants with clinical risk factors may slightly improve the predictive capacity for the future development of diabetes [25,26].

Adult obesity prevalence continues to rise according to new CDC data (Division of Nutrition Physical Activity and Obesity (DNPAO) September 15, 2021). By 2020, Adult Obesity Prevalence Maps showed that 16 states now have an adult obesity prevalence at or above 35% (Alabama, Arkansas, Delaware, Indiana, Iowa, Kansas, Kentucky, Louisiana, Michigan, Mississippi, Ohio, Oklahoma, South Carolina, Tennessee, Texas, and West Virginia). Combined data from 2018-2020 show notable racial and ethnic disparities. Among states and territories with sufficient data: 0 states had an obesity prevalence at or above 35% for non-Hispanic Asian residents, 7 states had an obesity prevalence at or above 35% for non-Hispanic White residents, 22 states had an obesity prevalence at or above 35% for Hispanic residents, 35 states and the District of Columbia had an obesity prevalence this high among non-Hispanic Black residents. Adults with obesity are at increased risk for many other serious health conditions such as heart disease, stroke, type 2 diabetes, some cancers and poorer mental health. Current evidence suggests that there are notable differences in the severity of psoriasis between racial and ethnic groups. Psoriasis affects the USA Latino population at a lower prevalence with more severe disease and a greater quality-of-life impact than their White counterparts with higher rates of co-morbidities, such as depression, obesity and diabetes [27].

In this study, we present significant evidence of association between *ADA*^*2*^ allele and obesity or BMIs greater than 25 in the Latino population. These data confirm that ADA may play a role in determining an individual’s BMI and therefore, risk for obesity. These results should add the *ADA*^*2*^ allele to the list of identified gene SNPs that are associated with obesity. Our findings of decreased *ADA*^*2*^ allele in obese subjects is consistent with the recent findings of Kumar et al. [28] demonstrating the antiglycemic, antihyperlipidemic, anti-inflammatory control and reduced ADA activity in DMT2 with garlic. Whether the reduced ADA activity resulted in increased adenosine systemically or in local subcellular micro-environments were not examined but may have consequences. Since obesity affects so many people in the world, it is essential that the causes of this disorder are identified so that its effects can be targeted and lifestyle can be modified. The current cohort should be expanded with a larger sample size of patients with a

BMI greater than 30. Identification of genetic markers could provide healthcare workers with the personalized medical information required to proactively treat or advise individuals with a genetic predisposition to obesity and medical conditions associated with it. Takhshid et al. [29] showed the SNP G22A in *ADA* was not associated with the risk of gestational diabetes mellitus (GDM) in the Iranian population, however the GG genotype was associated with poor glycemic control and obesity in GDM patients. Amoli et al. [30] observed a significant increase in the frequency of the ADA, AA genotype in obese Iranians compared to the controls plus significant increase in obese Iranian patients with triglyceride (TG) compared to the normal controls.

## Data Availability

All data produced in the present work are contained in the manuscript

## Abbreviations

ADA: adenosine deaminase
BMI: body mass index
DM1: type 1 diabetes mellitus
DM2: diabetes mellitus type 2
SNP: single nucleotide polymorphism
LDL-C: low density lipoprotein cholesterol
HDL-C: lower levels of high density lipoprotein cholesterol
DMSO: dimethylsulfoxide
dNTPs: deoxyribonucleotide triphosphates

## Acknowledgements

This research was supported in part by a grant from the CTSI grant-NCRR-NIH 1UL1RR029893.

